# Perceived public health threat a key factor for willingness to get the COVID-19 vaccine in Australia

**DOI:** 10.1101/2021.04.19.21255709

**Authors:** Rachael H Dodd, Kristen Pickles, Erin Cvejic, Samuel Cornell, Jennifer MJ Isautier, Tessa Copp, Brooke Nickel, Carissa Bonner, Carys Batcup, Danielle M Muscat, Julie Ayre, Kirsten J McCaffery

**Author notes:** Corresponding author: Dr Rachael Dodd, The University of Sydney, School of Public Health. Room 127A Edward Ford Building, (A27), The University of Sydney, NSW, 2006.

## Abstract

**Background:** Vaccination rollout against COVID-19 has begun across multiple countries worldwide. Although the vaccine is free, rollout might still be compromised by hesitancy or concerns about COVID-19 vaccines.

**Methods:** We conducted two online surveys of Australian adults in April (during national lockdown; convenience cross-sectional sample) and November (virtually no cases of COVID-19; nationally representative sample) 2020, prior to vaccine rollout. We asked about intentions to have a potential COVID-19 vaccine (If a COVID-19 vaccine becomes available, I will get it) and free-text responses (November only).

**Results:** After adjustment for differences in sample demographics, the estimated proportion agreeing to a COVID-19 vaccine if it became available in April (n=1146) was 76.2%. In November (n=2034) this was estimated at 71.4% of the sample; additional analyses identified that the variation was driven by differences in perceived public health threat between April and November. Across both surveys, female gender, being younger, having inadequate health literacy and lower education were associated with reluctance to be vaccinated against COVID-19. Lower perceived susceptibility to COVID-19, belief that data on the efficacy of vaccines is ‘largely made up’, having lower confidence in government, and lower perception of COVID-19 as a public health threat, were also associated with reluctance to be vaccinated against COVID-19. The top three reasons for agreeing to vaccinate (November only) were to protect myself and others, moral responsibility, and having no reason not to get it. For those who were indifferent or disagreeing to vaccinate, safety concerns were the top reason, followed by indecision and lack of trust in the vaccine respectively.

**CONCLUSIONS:** These findings highlight some factors related to willingness to accept a COVID-19 vaccine prior to one being available in Australia. Now that the vaccine is being offered, this study identifies key issues that can inform public health messaging to address vaccine hesitancy.

**HIGHLIGHTS:**

- Perceived public health threat is associated with intentions to vaccinate
- Those believing the efficacy of vaccines is made up were less willing to get vaccinated
- To protect myself and others was the top reason for getting the vaccine
- Safety concerns was the top reason against getting the vaccine

## INTRODUCTION

Rollout of nine variations of a vaccination against COVID-19 has now begun across ∼130 countries worldwide; including both high- and low-income countries. These countries are poised to ease restrictions implemented to prevent the spread of COVID-19 once the majority of their population has been vaccinated. As of April 15 2021, in the UK where a state of emergency was declared for COVID-19, over 30 million (∼45% of their population) have been partially vaccinated with a single dose of the Pfizer/BioNTech or AstraZeneca vaccine since December 2020.^1^ In Australia, where there are virtually no cases of COVID-19, 1.3 million (∼5% of the population) have been partially vaccinated since February 22 2021.^2^ Vaccines are crucial to developing herd immunity, protecting those who are most vulnerable to serious consequences of COVID-19, and to enable easing of national and international travel restrictions and opening up of the economy.

Willingness to get a COVID-19 vaccination (before it became available) has varied considerably across countries over the course of the pandemic. Between April and July 2020, willingness to vaccinate was shown to range from 57.6% in the US,^3^ to 64% in the UK^4^, 74% in New Zealand,^5^ and 85.8% in Australia.^6^ Our research in April showed inadequate health literacy and lower education were associated with a reluctance to be vaccinated^6^ and demonstrated an evident need to address health literacy, language and cultural needs of the community in public health messaging about COVID-19.^7^ Australian^8^ and New Zealand^5^ data have shown the most commonly reported reason to get vaccinated were to protect family and self, whilst safety about the vaccine was a chief concern.

Access to the vaccine is only one issue. Although the vaccine is free, rollout and uptake might still be compromised by concerns about COVID-19 vaccines, so it is important to investigate these. As our previous research during earlier months of the pandemic demonstrated high intentions towards vaccine uptake,^6,8^ it was important to reassess intentions since restrictions in Australia have been relaxed and the immediate threat of COVID-19 has diminished. This study aimed to examine vaccine willingness in the Australian population at two distinct time points in the pandemic: April 2020, during national lockdown, and November 2020, when there were virtually no cases of COVID-19 in Australia.

## METHODS

### Study Design

An online survey was conducted with two independent, cross-sectional samples at two different time points using the web-based survey platform Qualtrics. This study was approved by the University of Sydney Human Research Ethics Committee (2020/212).

### Setting

The survey was distributed Australia-wide. Data used for this study were collected between 17–22 April 2020, when national stage 3 restrictions (colloquially referred to as ‘lockdown’ at that time) had been in place for 3 weeks (i.e. only leaving home for essential reasons) and between 4-18 November 2020, when restrictions were considerably eased across Australia as no locally acquired cases were recorded for the first time since June 2020.

### Participants

Participants were aged 18 years and older, able to read and understand English, and currently residing in Australia. Participants were recruited via Dynata, who have more than 600 000 online Australian panel members aged older than 18 years who have consented to participate in online research. Panel members were sent an email invitation to participate in the study and received points for completing the survey, which they could redeem for gift vouchers, donations to charities, or money. As the April sample was overrepresented by those having attended University, we purposively set quotas to recruit a nationally representative sample by age, gender and education in the November sample.

### Measures

Participants completed sociodemographic questions of age, gender and educational status. We assessed health literacy using the Single Item Literacy Screener (SILS).^9^ Participants rated their perceived risk of contracting COVID-19, ‘Do you think that you will get sick from COVID-19?’ (not at all/it’s possible/I probably will/I definitely will) and the perceived public health threat of COVID-19, ‘On a scale of 1 to 10, how serious of a public health threat do you think COVID-19 is currently’. Participants were also asked whether they have trust in institutions (scientists involved in developing and testing new ways to control COVID-19, researchers involved in tracking and predicting COVID-19 cases, and medical institutions (GPs, hospitals) involved in managing COVID-19 cases) with the responses on a seven-point Likert scale from 1 (do not trust at all) to 7 (trust very much). Confidence in the federal government was measured, ‘How confident are you that the federal government can prevent further outbreak of COVID-19?’. Two items were included assessing agreement with misinformation (‘Data about the effectiveness of vaccines is often made up’ and ‘The threat of COVID-19 is greatly exaggerated’ (seven-point scale: strongly disagree to strongly agree).

Participants were asked to respond on a seven-point Likert scale about intentions to have a potential COVID-19 vaccine (If a COVID-19 vaccine becomes available, I will get it). In November, the participants were also asked to give a reason for their choice (free-text response). Details of all measures are included in our baseline survey paper on health literacy disparities.^7^

### Statistical Analysis

Statistical analyses were conducted using Stata/IC v16.1 (StataCorp, College Station, TX). Descriptive statistics were generated for demographic characteristics, COVID-19 beliefs, and willingness to get a COVID-19 vaccine, and compared between cross-sectional (April and November) samples using independent t-tests for continuous variables and chi-square tests for categorical variables. Differences in willingness to get a COVID-19 vaccine between samples were examined using ordered logistic regression, controlling for age group, gender, education, and health literacy adequacy (base model). A series of exploratory analyses using ordered logistic regression were then conducted to identify potential factors associated with vaccine willingness by collectively adding personal risk belief, vaccine efficacy beliefs, confidence in government, institutional trust, and perceived public health threat into the base model (full model). Participants who responded strongly agree, agree or somewhat agree were coded as ‘agree’; strongly disagree, disagree, somewhat disagree as ‘disagree’; and neither agree nor disagree was standalone.

Free-text responses were analysed using content analysis,^10^ a widely used analysis method which combines qualitative and quantitative methods to analyse text data, allowing the content and frequency of categories to be reported. Members of the research team (RD, KP) first read through all the free-text responses (n=2034), removed responses with no comments (n=145), leaving 1889 free text responses, and developed the initial coding framework. Three members of the research team (SC, JI, TC) reviewed the free-text responses and discussed the initial coding framework. A random selection (randomised in Microsoft Excel) of 100 responses were triple coded independently by three members of the research team (SC, TC & JI). Level of agreement was tested using Fleiss Kappa^11^ and indicated substantial agreement (κ=0.756). Any discrepancies were discussed between SC, TC & JI until consensus was reached. The coding framework was then amended to include extra themes. SC, TC and JI then independently coded approximately 600 responses each. The frequency of each code and main themes were then reported.

## RESULTS

Sample characteristics are displayed in Table 1. Respondents sampled in November were selected to be nationally representative of the Australian population (by age, gender and education), and as such were younger (p<.001) and had a more even distribution of education level compared to the April sample which had a high proportion of university-educated adults (p<.001). A slightly greater proportion of the November sample had adequate health literacy compared to the April sample (p=.015). Relative to the group sampled in April, a greater proportion of the November sample believed they were unlikely to get sick with COVID-19 (p=.002), that data about the efficacy of vaccines (in general) is made up (p=.002), perceived the general public health threat of COVID-19 to be lower (p<.001), and that the threat of COVID-19 is greatly exaggerated (p<.001). Willingness to get a COVID-19 vaccine if available was also lower in November compared to April (p<.001).

**Table 1.**
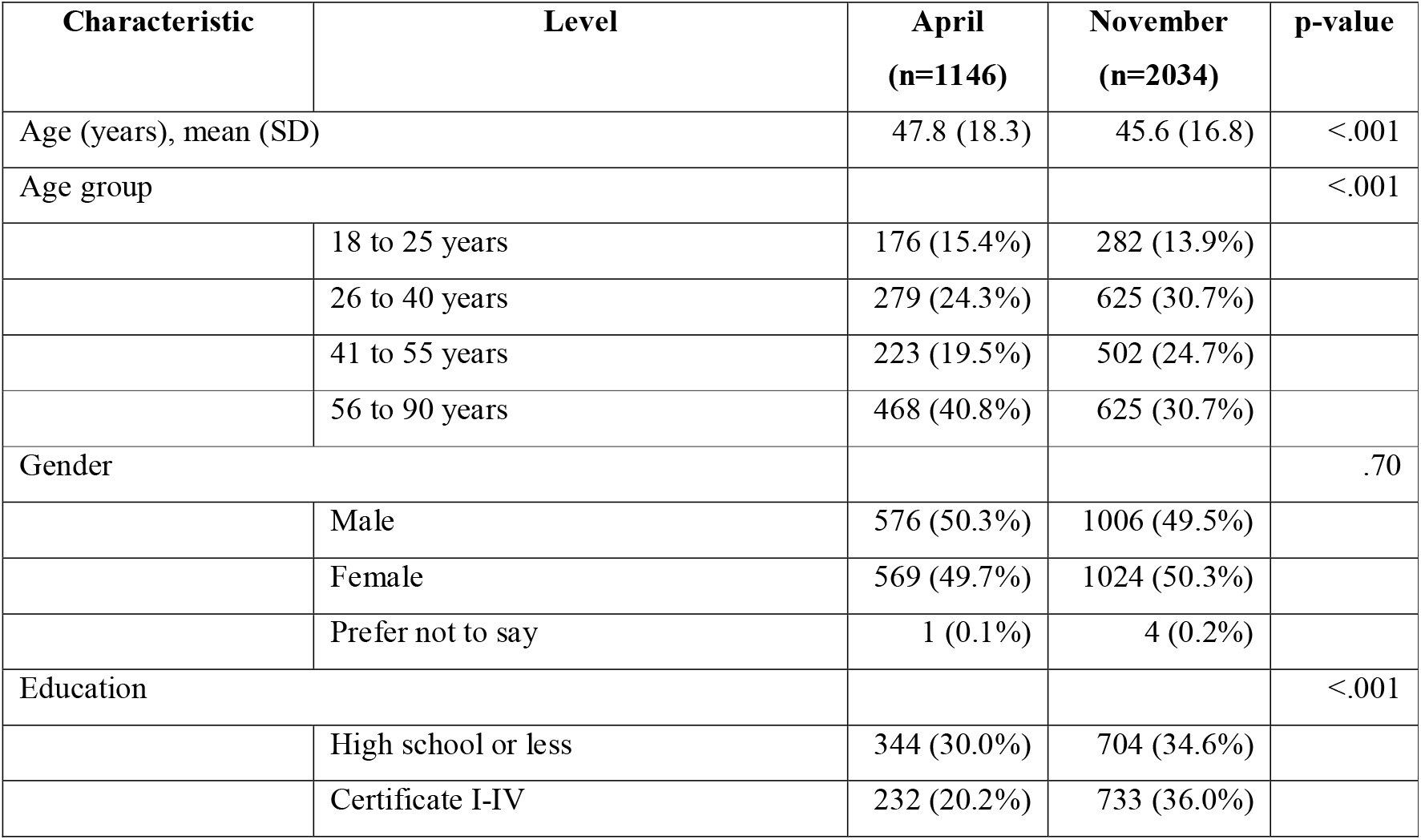
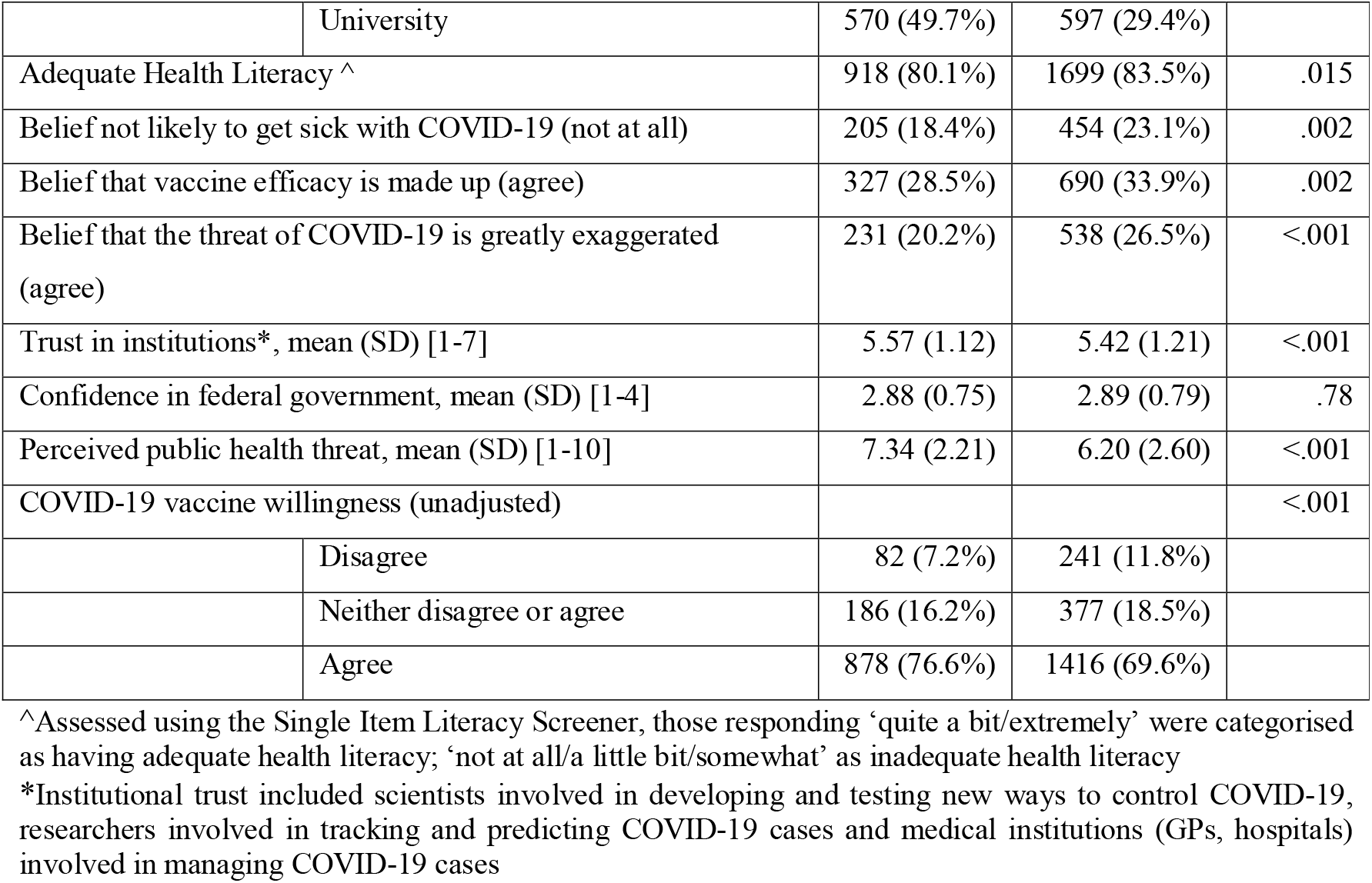
Sample characteristics of cross-sectional Australian samples collected in April 2020 (n=1146) and November 2020 (n=2034). Data are displayed as n (%) unless indicated otherwise.

| Characteristic | Level | April<br>($n=1146$) | November<br>($n=2034$) | p-value |
| --- | --- | --- | --- | --- |
| Age (years), mean (SD) |  | 47.8 (18.3) | 45.6 (16.8) | <.001 |
| Age group |  |  |  | <.001 |
|  | 18 to 25 years | 176 (15.4%) | 282 (13.9%) |  |
|  | 26 to 40 years | 279 (24.3%) | 625 (30.7%) |  |
|  | 41 to 55 years | 223 (19.5%) | 502 (24.7%) |  |
|  | 56 to 90 years | 468 (40.8%) | 625 (30.7%) |  |
| Gender |  |  |  | .70 |
|  | Male | 576 (50.3%) | 1006 (49.5%) |  |
|  | Female | 569 (49.7%) | 1024 (50.3%) |  |
|  | Prefer not to say | 1 (0.1%) | 4 (0.2%) |  |
| Education |  |  |  | <.001 |
|  | High school or less | 344 (30.0%) | 704 (34.6%) |  |
|  | Certificate I-IV | 232 (20.2%) | 733 (36.0%) |  |
|  | University | 570 (49.7%) | 597 (29.4%) |  |
| Adequate Health Literacy ^ |  | 918 (80.1%) | 1699 (83.5%) | .015 |
| Belief not likely to get sick with COVID-19 (not at all) |  | 205 (18.4%) | 454 (23.1%) | .002 |
| Belief that vaccine efficacy is made up (agree) |  | 327 (28.5%) | 690 (33.9%) | .002 |
| Belief that the threat of COVID-19 is greatly exaggerated (agree) |  | 231 (20.2%) | 538 (26.5%) | <.001 |
| Trust in institutions*, mean (SD) [1-7] |  | 5.57 (1.12) | 5.42 (1.21) | <.001 |
| Confidence in federal government, mean (SD) [1-4] |  | 2.88 (0.75) | 2.89 (0.79) | .78 |
| Perceived public health threat, mean (SD) [1-10] |  | 7.34 (2.21) | 6.20 (2.60) | <.001 |
| COVID-19 vaccine willingness (unadjusted) |  |  |  | <.001 |
|  | Disagree | 82 (7.2%) | 241 (11.8%) |  |
|  | Neither disagree or agree | 186 (16.2%) | 377 (18.5%) |  |
|  | Agree | 878 (76.6%) | 1416 (69.6%) |  |

^Assessed using the Single Item Literacy Screener, those responding 'quite a bit/extremely' were categorised as having adequate health literacy; 'not at all/a little bit/somewhat' as inadequate health literacy
\*Institutional trust included scientists involved in developing and testing new ways to control COVID-19, researchers involved in tracking and predicting COVID-19 cases and medical institutions (GPs, hospitals) involved in managing COVID-19 cases

### Differences in vaccine willingness in April and November

After adjustment for age group, gender, education, and health literacy adequacy, there was evidence of a difference in vaccine willingness between samples (Table 2). Predicted probabilities (estimated at the sample means for all covariate values) suggested a lower proportion of individuals being willing to agree to a vaccine in November (71.4%) compared to April (76.2%), representing an absolute difference of 4.8% (95%CI: 1.5 to 8.0%; p=.004; see Figure 1). The adjusted odds of having a higher level of willingness to get a COVID-19 vaccine in the November sample was 0.78 (95% CI: 0.66, 0.93; p=.005) times that of the April sample.

**Table 2.** Results from ordered logistic regression of willingness to get a COVID-19 vaccine, controlling for demographic variables. The outcome was coded as disagree, neither disagree or agree, and agree. Values are provided as adjusted odds ratios (aOR) with corresponding 95% confidence intervals.

| Variable | Level | aOR (95% CI) | p-value |
| --- | --- | --- | --- |
| Time Point |  |  | .005 |
|  | April | 1.00 (ref) |  |
|  | November | 0.78 (0.66, 0.94) |  |
| Age group (vs 18-25 years) |  |  | <.001 |
|  | 26-40 years | 0.90 (0.70, 1.14) |  |
|  | 41-55 years | 0.94 (0.73, 1.21) |  |
|  | 56-90 years | 1.78 (1.37, 2.31) |  |
| Female Gender (vs Male)* |  | 0.79 (0.67, 0.93) | .005 |
| Education (vs High school or less) |  |  | <.001 |
|  | Certificate I-IV | 1.04 (0.85, 1.25) |  |
|  | University | 1.77 (1.45, 2.17) |  |
| Adequate health literacy (vs inadequate) |  | 1.63 (1.34, 1.98) | <.001 |

\*Gender = “Other / Prefer not to say” was included in the model, however results are not displayed due to likely instability of estimates owing to the small sample size in this group (n=5).

**Figure 1.**
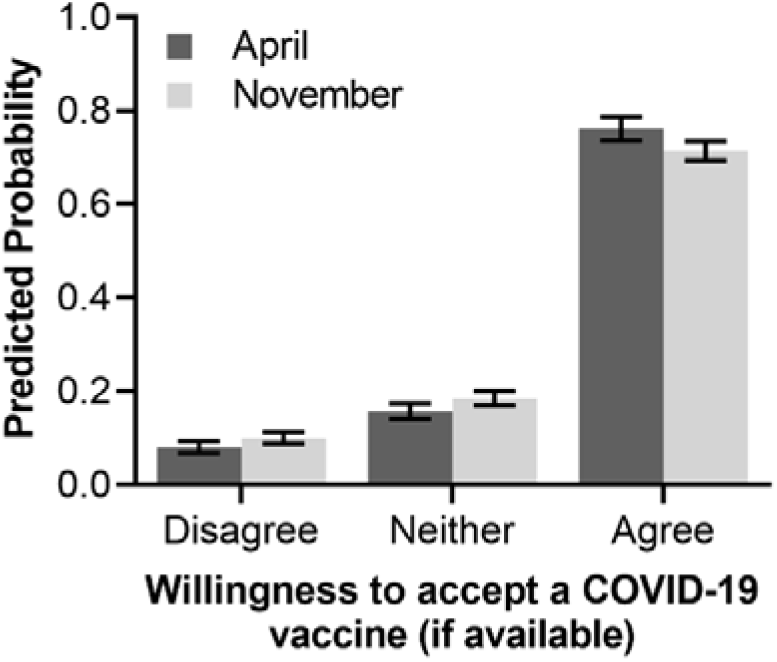
Predicted probabilities of willingness to get a COVID-19 vaccine (if available) by cross-sectional survey time point (April vs November) after adjustment for age, gender, education, and health literacy adequacy. Estimates were determined at covariate sample means. Error bars indicate the 95% confidence interval.

### Factors associated with COVID-19 vaccine willingness

The results from the full ordered logistic regression model are shown in Table 3 for both time points. Older age (i.e., individuals aged 56 to 90 years, relative to all younger age groups), university education (relative to high school education or less), adequate health literacy, higher confidence in government, trust in institutions, and greater perception of COVID-19 as a public health threat were associated with increased odds of being more willing for vaccination (ie in a higher vaccine willingness category). Female gender (relative to male gender), a low personal perceived risk of COVID-19, and belief that the data on efficacy of vaccines is largely made up were associated with reduced odds of being in a higher vaccine willingness category. After controlling for these factors, there was no longer statistical evidence of a difference between timepoint samples in willingness to vaccinate (aOR = 0.98, 95%CI: 0.81, 1.17; p=.80).

**Table 3.** Results from multivariable ordered logistic regression model of willingness to get a COVID-19 vaccine. The outcome was coded as disagree, neither disagree or agree, and agree. Values are provided as adjusted odds ratios (aOR) with corresponding 95% confidence intervals.

| Variable | Level | aOR (95% CI) | p-value |
| --- | --- | --- | --- |
| Time Point |  |  | .72 |
|  | April | 1.00 (ref) |  |
|  | November | 0.97 (0.80, 1.17) |  |
| Age group (vs 18-25 years) |  |  | <.001 |
|  | 26-40 years | 0.75 (0.58, 0.98) |  |
|  | 41-55 years | 0.81 (0.61, 1.06) |  |
|  | 56-90 years | 1.36 (1.02, 1.80) |  |
| Female Gender (vs Male)* |  | 0.69 (0.57, 0.82) | <.001 |
| Education (vs high school or less) |  |  | <.001 |
|  | Certificate I-IV | 1.03 (0.84, 1.27) |  |
|  | University | 1.53 (1.23, 1.90) |  |
| Adequate health literacy (vs inadequate) |  | 1.15 (0.93, 1.43) | .19 |
| Belief not likely to get sick with COVID-19 (vs likely) |  | 0.58 (0.47, 0.71) | <.001 |
| Belief that vaccine efficacy is made up (vs disagree) |  | 0.83 (0.69, 0.99) | .039 |
| Confidence in federal government (/ unit) |  | 1.15 (1.02, 1.29) | .023 |
| Trust in institutions (/unit) |  | 1.75 (1.62, 1.90) | <.001 |
| Perceived public health threat (/unit) |  | 1.16 (1.12, 1.20) | <.001 |

\*Gender = “Other / Prefer not to say” was included in the model, however results are not displayed due to likely instability of estimates owing to the small sample size in this group (n=5).

To better understand which (if any) of the additional covariates entered into the model were accounting for the differences between timepoints observed from the base model, a leave-one-out approach was employed whereby the additional covariates were individually removed from the model and the resulting coefficient for timepoint compared to the full model. Model coefficients remained consistent using this backwards removal approach, except when perceived public health threat of COVID-19 was removed. When this covariate as omitted, there was statistical evidence (p=.043) of lowered odds of higher vaccine willingness in November (aOR: 0.83, 95%CI: 0.69, 0.99) compared to April. Thus, it appears differences in the perceived public health threat between April and November were driving the observed difference in vaccine willingness (see Figure 2).

**Figure 2.**
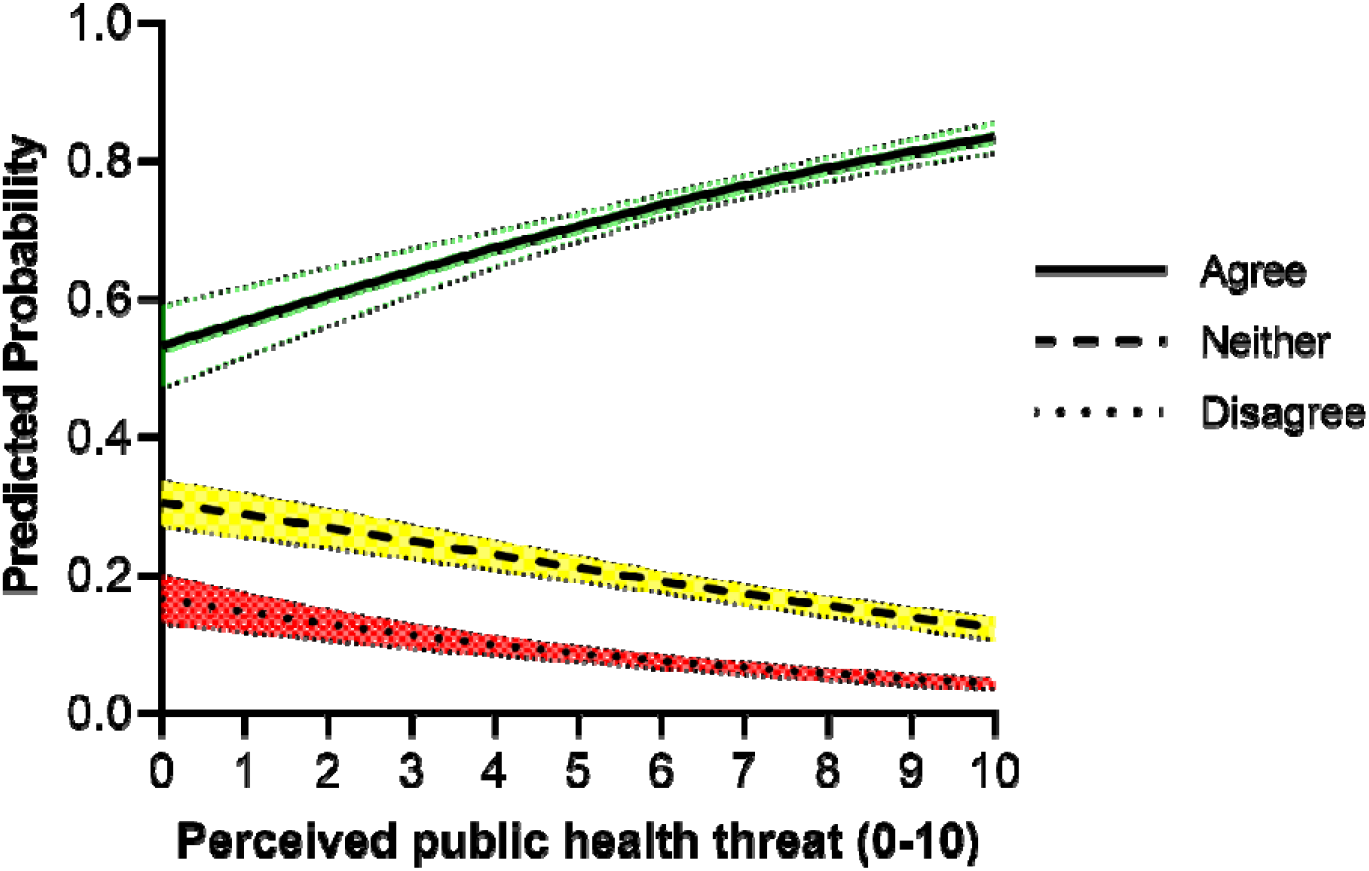
Predicted probabilities of willingness to get a COVID-19 vaccine (if available) by perceived public health threat. Values are estimated at the mean value of all other model covariates. Shaded bands indicate the 95% confidence interval.

### Content Analysis Results

Of the 2034 participants who provided a response to the question (‘If a COVID-19 vaccine becomes available, I will get it’) in November, 1889 (93%) provided a written response explaining their choice. A total of 41 themes were generated to capture responses to the question, with 20 ‘agree’ themes and 21 ‘disagree’ themes.

Table 4 (full table Appendix 1) presents results of a content analysis showing the most common reasons for willingness or reluctance to get a COVID-19 vaccine, including example free-text responses. The top three reasons for agreeing to vaccinate were: 1) to *protect myself and others* (23.5%, n=326/1388), 2) *moral responsibility* (10.4%, n=145/1388), and 3) *no reason not to get it* (9.5%, n=132/1388), expressing a ‘nothing to lose’ attitude with getting the vaccine. The top reasons for being reluctant to have the vaccine were: 1) concern about the *safety of the vaccine* (25.3%, n=61/241), 2) *lack of trust* in the vaccine (20.7%, n=50/241), and 3) *simply disagreeing* (16.2%, n=39/241). For those who neither agreed or disagreed that they would have the vaccine, reasons give were: 1) *safety concerns* (29.8%, n=116/389), 2) being *undecided* (23.1%, n=90/389), and 3) *needing more information* (19%, n=74/389).

**Table 4.** Themes identified in free-text responses (n=1889) with example responses.

|  | N | % | Example Free Text Response |
| --- | --- | --- | --- |
| **Agree (n=1388)\*** |  |  |  |
| To protect myself and others | 326 | 23.5 | "Helps to protect others and my family" |
| Moral responsibility | 145 | 10.4 | "I think it is the sensible thing to do for the good of myself, family and friends, and society in general." |
| No reason not to get it | 132 | 9.5 | "It can't hurt" |
| To stop the virus | 125 | 9.0 | "Immunisation is the only way to control covid-19" |
| Depends on proven safety | 115 | 8.3 | "If it has been suitably tested I will be happy to have it." |
| Simply agree | 113 | 8.1 | "I agree" |
| **Disagree (n=241)\*** |  |  |  |
| Safety concerns | 61 | 25.3 | "Don't feel comfortable putting that kind of thing in my body" |
| Vaccine or government trust | 50 | 20.7 | "I have a weird feeling about it" |
| Simply disagree | 39 | 16.2 | "Don't want to" |
| Other | 22 | 9.1 | "Do not like" |
| Need more information | 19 | 7.9 | "Don't know enough about it" |
| Not at risk | 17 | 7.1 | "Where I am there was 1 case since this started and he came from overseas. I don't feel I have been put at risk of getting it so why put that into my body when there is no need at this time." |
| **Neither agree nor disagree (n=389)\*** |  |  |  |
| Safety Concerns | 116 | 29.8 | "Has to be proven safe" |
| Undecided | 90 | 23.1 | "Definitely undecided" |
| Need more information | 74 | 19.0 | "I need more information about its safety condition before I use it" |
| Other | 29 | 7.5 | "Only if made in Australia" |
| Responses not related to survey question | 14 | 3.6 | "I can see into the future" |

\*some free-text responses were allocated more than one code

## DISCUSSION

This Australian data from two cross-sectional samples at two time points during the COVID-19 pandemic demonstrates that willingness to vaccinate against COVID-19 is predominately driven by the perceived public health threat of COVID-19 in Australia. In the November sample, when cases were low, restrictions were eased and life was much more ‘normal’, vaccine willingness was lower than in the April sample, during national lockdown. We found that 18-23% of respondents across the samples believed the threat of COVID-19 is exaggerated and 29-34% believed the effectiveness of vaccines (in general) is made up.

Being female, having a belief that oneself is not likely to get COVID-19, and belief that the data on efficacy of vaccines is largely made up were associated with being less willing to have the COVID-19 vaccine. Older age (55+ years), university educated, adequate health literacy, higher confidence in government, and greater perception of COVID-19 as a public health threat were associated with greater willingness to be vaccinated against COVID-19. The top reason given for being willing to have the COVID-19 vaccine was to protect myself and others, with concern about the safety of the vaccine being the primary reason for being less willing to have the COVID-19 vaccine.

Our findings showed that perceived public health threat of COVID-19 was associated with intentions to vaccinate, and belief that the threat of COVID-19 is exaggerated was associated with less intention to vaccinate, in both samples. If people are less likely to have a COVID-19 vaccine due to reduced perception of public health threat in Australia, then this could potentially threaten the government aim of achieving 95% uptake of the COVID-19 vaccine.^12^

We know from previous infectious disease outbreaks that perceived risk is influential in people taking preventative measures,^13^ and people also need to believe that the behaviour, in this case having the vaccine, will be effective in reducing their risk.^14^ Previous research has unsurprisingly shown that having personal or direct experience with COVID-19 is associated with greater perceived risk, and perceived risk during the COVID-19 pandemic has been highest in the UK compared to US, Australia, Germany, Spain, Italy, Sweden, Mexico, Japan and South Korea.^15^ Given the association between believing vaccine efficacy is made up and reduced intention to vaccinate in our study, there is a need for strategies that reduce people’s complacency about the public health threat of COVID-19, as well as correct misperceptions about vaccine efficacy, safety and importance. Strategies could also aim to communicate the cost of not vaccinating to society rather than the individual, particularly for those younger age groups who perceive themselves at less risk and are therefore less willing to get the vaccine. Previous research has shown that people’s prior beliefs are influential in trusting facts, but not in response to communication of uncertainty.^16^ This is encouraging as this means being transparent about the uncertainties about the vaccine should not undermine people’s trust in the facts or who is communicating these.

Some recommendations from the National Centre for Immunisation Research and Surveillance, based on a review of COVID-19 acceptance literature, include addressing doubts about the pandemic threat by explaining complex concepts in ways that are easy to understand, and addressing low perceived risk by emphasising the broad range of benefits of the vaccine.^17^ Similar strategies to those used for getting tested for COVID-19 could also be adapted in public communication, for example using celebrities in television advertisements. Findings from our previous national surveys have concluded that any communication needs to be designed for those with lower health literacy and education and appropriate for culturally and linguistically diverse groups and Aboriginal and Torres Strait Islander people.^7,8^

Safety concerns were the top reason for participants to be indifferent or disagree with having the vaccine. Since this data was collected, concerns have been raised in the early stages of the vaccine rollout internationally about whether the AstraZeneca vaccine is implicated in thromboembolic events, and some European countries temporarily suspended the use of the vaccine while these events were investigated.^18^ In Australia, Pfizer is now the recommended vaccine for those under 50 years of age due to the potentially increased risk following AstraZeneca vaccine in this age group.^19^ Although the likelihood of these events occurring is low, suspension of the AstraZeneca vaccine and a change in guidelines has likely impacted public confidence globally and it often takes time for this to recover.^20^ Safety concerns were the top reason we found for those unwilling to be vaccinated, so it is of upmost importance to restore public confidence through being transparent about the decisions made and the data collected monitoring the side effects, particularly through the AusVax program.^21^ Those chosen to communicate about these issues need to do so with empathy, and not dismiss concerns about the vaccine.^20^ Trust in the federal government and in doctors is high in Australia.^6^ Given GPs and other health professionals will play a key role in administering the vaccine, restoring public confidence and alleviating concerns may fall to them, so ensuring strong communication channels between doctors and the government is vital.

Our finding that women have lower intentions to be vaccinated than men, has been found in previous research in Australia^22^ and in 35/60 studies in a recent systematic review looking at gender differences in intentions to vaccinate against COVID-19.^23^ Potential reasons for this finding could be due to women who are pregnant, breastfeeding, or planning children being concerned about vaccine safety, although these reasons were not evident in the free text responses of the current study. Due to the exclusion of pregnant women in the large vaccine trials,^24^ there is no recommendations about vaccinating women who are pregnant, breastfeeding or planning a pregnancy, but decision aids are available to help women make an informed choice about the COVID-19 vaccine.^25^ It is pertinent that we conduct further research with younger women to understand their reasons for vaccine hesitancy.

The current study is strengthened by presenting findings from two large cross-sectional samples. Participants were only asked for their reasons behind their intentions in the November survey and therefore we cannot draw any conclusions on whether these reasons have changed over the course of the pandemic. This survey measured participants intentions to have the COVID-19 vaccine in the future and not actual behaviour and therefore further research since the rollout of the vaccine is required.

These findings show that perceived public health threat is an important factor in driving vaccine intentions. Understandably in Australia where incidence of COVID-19 is virtually none, perceptions of public health threat have decreased during the pandemic. Public health messaging, from both the government and trusted health professionals, must aim to reduce people’s complacency about the risk of COVID-19 and their belief that efficacy of vaccines is often made up, particularly if our international borders are to open in the future.

## Data Availability

Study data may be made available on request to accredited researchers who gain ethical approval.

## Acknowledgements

We thank Dynata for recruitment services and all study participants.

