## Supplementary Tables 1-3 for "Perceived public health threat a key factor for willingness to get the COVID-19 vaccine in Australia"

**APPENDIX**

Table S1: ‘Agree’ themes identified in free-text responses (n=1388) with example response.

| **Agree** | **N** | **%** | **Example Free Text Response** |
| --- | --- | --- | --- |
| To protect myself and others | 326 | 23.5 | “Helps to protect others and my family” |
| Moral responsibility | 145 | 10.4 | “I think it is the sensible thing to do for the good of myself, family and friends, and society in general.” |
| Why not? | 132 | 9.5 | “It can't hurt and would give a certain amount of feeling safer” |
| To stop the virus | 125 | 9.0 | “immunisation is the only way to control covid-19” |
| Depends on proven safety | 115 | 8.3 | “If it has been suitably tested I will be happy to have it.” |
| Simply agree | 113 | 8.1 | “I agree” |
| Vaccine or government trust | 56 | 4.0 | “The past has shown how vaccines have helped eradicate viruses” |
| High risk health wise | 52 | 3.7 | “I am in the high risk category, so what do I have to lose?” |
| To travel | 49 | 3.5 | “It will make me feel more comfortable about travelling.” |
| Nonsensical | 44 | 3.2 | “a psychic said not to get it if the vaccine is white. if it is, I won’t be having it.” |
| To get back to normal | 40 | 2.9 | “I think it will help me to be able to live my life pre COVID-19” |
| Other | 38 | 2.7 | “It will depend on how Australia is currently going with COVID when and if they ever create a vaccine” |
| I would wait | 36 | 2.6 | “Because this is a new vaccine I want to wait a month or so to see if there are any adverse reactions” |
| If it is available to me | 36 | 2.6 | “I will get it if it becomes available!!!!!!” |
| More information required before getting it | 23 | 1.7 | “Depends on what when where type of vaccine” |
| Recommendation from doctor or government | 23 | 1.7 | “I will wait until my Doctor tells me too.” |
| Not sure | 18 | 1.3 | “not really sure” |
| Required for my job | 13 | 0.9 | “probably be mandatory for working with the general public” |
| Social pressure | 2 | 0.1 | “I would feel judged if I didn’t” |
| To protect the economy | 2 | 0.1 | “I believe it’s a very dangerous evolving virus that we need protection from so we can repair economy and employment problems” |

Table S2: ‘Disagree’ themes identified in free-text responses (n=241) with example response.

| **Disagree** | **N** | **%** | **Free text comment** |
| --- | --- | --- | --- |
| Safety concerns | 61 | 25.3 | “Don’t feel comfortable putting that kind of thing in my body” |
| Vaccine or government trust | 50 | 20.7 | “I have a weird feeling about it” |
| Simply disagree | 39 | 16.2 | “Don’t want to” |
| Other | 22 | 9.1 | “It all depends on when it is found” |
| Need more information | 19 | 7.9 | “Don’t know enough about” |
| Not at risk | 17 | 7.1 | “Where I am there was 1 case since this started and he came from overseas. I don't feel I have been put at risk of getting it so why put that into my body when there is no need at this time.” |
| Comorbidities | 7 | 2.9 | “I am allergic to flu vaccines” |
| Undecided | 6 | 2.5 | “Don’t know” |
| Nonsensical response | 6 | 2.5 | “This vaccine will not allow us to experience our great solar flash!” |
| Rather get COVID/ herd immunity | 4 | 1.7 | “I'd rather take the chance and develop a natural immunity due to exposure when the time comes. Also my understanding is vaccines often have a slight negative affect on health for a long period of time often mild flu like symptoms.” |
| High risk first | 3 | 1.2 | “I think the COVID-19 should be made available to those most vulnerable in the community first and then the general public so I won't get vaccinated until those at high risk are.” |
| COVID-19 is a hoax | 2 | 0.8 | “I don't believe in COVID19” |
| I don’t like vaccines/needles | 2 | 0.8 | “I don't like VACCINES” |
| Only if recommended by my doctor | 1 | 0.4 | “I will wait side effects are of great concern and as a result I will delay having any kind of vaccine for con vid 19 until I have been given the ok to do so by my gp” |
| If necessary to travel | 1 | 0.4 | “I will only get it if I need to get it to go overseas travelling or something - which I am dying to do.” |
| If it is mandatory | 1 | 0.4 | if mandatory |

Table S3: ‘Neither agree nor disagree’ themes identified in free-text responses (n=389) with example response.

| **Neither agree nor disagree** | **N** | **%** | **Example free text response** |
| --- | --- | --- | --- |
| Safety Concerns | 116 | 29.8 | “Has to be proven safe” |
| Undecided | 90 | 23.1 | “Definitely undecided” |
| Need more information | 74 | 19.0 | “I need more information about it's safety condition before I use it” |
| Other | 29 | 7.5 | “only if made in Australia” |
| Nonsensical response | 14 | 3.6 | “I can see into the future” |
| Only if recommended by my doctor | 13 | 3.3 | “I will do my research first and ask healthcare professionals I trust” |
| Not at risk | 13 | 3.3 | “I don't think I'll ever get COVID” |
| Vaccine or government trust | 9 | 2.3 | “don’t know if I trust it” |
| Simply disagree | 9 | 2.3 | “No particular reason” |
| If it is mandatory | 8 | 2.1 | “Only if I have to.” |
| I don’t like vaccines/needles | 6 | 1.5 | “I hate needles” |
| High risk first | 3 | 0.8 | “others may have priority” |
| Cost | 2 | 0.5 | “Probably, if they do find one and if it's reasonably priced.” |
| COVID-19 is a hoax | 1 | 0.3 | “covid is fake” |
| If necessary to travel | 1 | 0.3 | “I will only get it, if it is necessary to leave the country” |
| Comorbidities | 1 | 0.3 | “I’m immunodeficient” |
